# The Impacts of Neighborhood Disparities on US Population Health During the COVID-19 Pandemic: A Literature Review and Policy Analysis for Future Response

**DOI:** 10.1101/2024.09.12.24313566

**Authors:** Brianna M. White, Whitney S. Brakefield, Olufunto A. Olusanya, Rameshwari Prasad, Fekede Kumsa, Soheil Hashtarkhani, Arash Shaban-Nejad

## Abstract

**Introduction/Background:** The COVID-19 pandemic has disrupted major aspects of daily life since its emergence in late 2019, impacting health and well-being worldwide. Aside from genetic predisposition, numerous external factors play key roles in determining levels of disease susceptibility and overall health status.

**Aim/Objectives:** This review and policy analysis sought to examine literature focused on the impact of neighborhood environments on population health and well-being during the COVID-19 pandemic period. Examined studies explore the impacts of various aspects of the neighborhood environment (i.e., walkability, transportation, access to healthcare/vaccination, etc.) during the COVID-19 era on population health and well-being.

**Methods:** Databases were searched for studies published from March 2020 to August 2022. Selected studies examine the impacts of various aspects of the neighborhood environment (i.e., walkability, transportation, access to healthcare/vaccination, climate, crime rates, persistent poverty, the social spread of misinformation, etc.) during the COVID-19 era on population health and well-being.

**Results:** Findings from our review suggest that those living in less-than-favorable environments were faced with significant challenges, making vulnerable populations more prone to the severity of COVID-19 infection.

**Conclusion:** We argue these results establish cause for the consideration of the effects of neighborhood environments in developing intelligent intervention strategies for both pandemic recovery and preparation for future public health emergencies, especially among vulnerable populations.

## 1. Introduction

The emergence of COVID-19 in late 2019 disrupted daily life worldwide, causing physiological, psychological, and psychosocial consequences on both individual and population levels.^1^ Since the World Health Organization (WHO) declared a public health emergency in response to COVID-19 in early 2020, preventive measures such as face mask mandates, social distancing, and hand hygiene practices have been implemented to curb the viral spread.^1^ Despite the best efforts, more than 100 million confirmed cases of infection and over 1 million deaths were documented in the United States (US).^2^ Apart from genetic predisposition, socio-ecological models explain many environmental stressors determine the levels of illness susceptibility and overall health status.^3^ External factors such as occupation and other societal patterns have also been identified as major determinants in COVID-19 infection outcomes.^4,5^ Overall, the neighborhoods in which people live have major impacts on overall health and well-being, so much so that Healthy People 2030 has given high priority to improving unfit neighborhood conditions.^6^

The relationship between neighborhood environments and well-being has been extensively researched and documented^7, 8^, with strong evidence indicating that individuals living in deprived and unequal neighborhoods tend to have lower levels of subjective well-being compared to those without compounding challenges.^9,10^ As communities implemented COVID-19 mitigation measures, neighborhood inequities became more evident. Moreover, communities may have been forced to confront pre-existing spatial inequities, which were further exacerbated by the often-crowded residential areas within these communities. As a result of the restriction on movement, populations were not able to move about as frequently as they did before the pandemic.^11^ Concerning the effects of COVID-19, multiple studies have examined the variation in morbidity and mortality rates between communities of different backgrounds in relation to their population density, income, and access to healthcare or insurance coverage.^12, 13, 14^ While research is ongoing, there is a need to further explore the effects of various neighborhood environment characteristics on access to routine healthcare, including preventive services such as vaccination, during the pandemic period.

Attention to inequalities related to neighborhood environments and how neighborhood characteristics have influenced population health could inform innovative strategies to tackle growing disparities, spark the development of effective policy solutions, and aid in equitable preparation for future outbreaks. In this article, we undertake a comprehensive analysis of currently available literature examining how factors in the neighborhood environment impact population health outcomes during the COVID-19 pandemic. We argue our findings could provide a better understanding of how these characteristics have influenced population health, informing innovative strategies to tackle neighborhood disparities amid the ongoing COVID-19 crisis and aid in equitable preparation for future outbreaks.

## 2. Methods

### 2.1. Search Strategy

The review of literature incorporated studies that were identified from three databases (PubMed, Social Science Research Network (SSRN), and Google Scholar) published between March 2020 to August 2022. We adopted March 2020 to the present as our timeline for study selection because we felt this represented literature published during the COVID-19 pandemic era. Our search terms were combined and utilized based on the following thesaurus and keywords: “Neighborhood environments”, “Built neighborhood environment”, “Biophysical neighborhood environment”, “Social neighborhood environment”, “Vaccination uptake”, “Vaccination hesitancy”, “Persistent poverty”, “Social Determinants of Health”, “Social Determinants of COVID-19”, “SARS-CoV-2”, “COVID-19”, “Neighborhood”, “Household composition”, “Income/Income Level”, “Crime”, “Transportation Disparities”, and “Housing Disparities”. Search terms and keywords were combined to search phrases including: “Neighborhood environments and vaccination uptake”, “Neighborhood environments and routine childhood vaccination”, “Social Determinants of Health and COVID-19”, “Persistent poverty and COVID-19”, “Neighborhood environment and persistent poverty”, “Effects of neighborhood environment on vaccination uptake”, “Effects of built neighborhood environment on vaccination uptake”, “Effects of biophysical neighborhood environment on vaccination uptake”, “Effects of social neighborhood environment on vaccination uptake”, “Education and/or Income Level and vaccination hesitancy”, “Crime rates and vaccination uptake/hesitancy”, “Violent Crime and vaccination uptake/hesitancy”, “Transportation and Health Disparities, vaccination uptake/hesitancy”, and “Housing and/or Neighborhood and Health Disparities, vaccination uptake/hesitancy”.

### 2.2. Eligibility Criteria

Studies were included for review if they met the following eligibility criteria: (1) examined elements of neighborhood environments – either built (walkability, food environments), biophysical (air pollution, green space, climate), or social (deprivation, safety, social cohesion, politics, mistrust/misinformation) - barriers/access to vaccination, etc.) as predictor variables, (2) were conducted in the U.S., and (3) were available in English language. Studies were excluded if they neither focused on the effects of neighborhood environments on population health nor vaccination uptake/hesitancy. Abstracts, reviews, letters, editorials, and theoretical papers were also excluded.

### 2.3. Study Selection, Data Extraction, and Analysis

The initial electronic database search generated 562 articles whose titles/abstracts were screened by reviewers (BW and RP) to ensure fit to the specified eligibility criteria. 475 studies were eliminated because they did not meet the inclusion criteria or were duplicates. After title/abstract screening and full-text review, a total of 87 studies were selected for data extraction and analysis, as demonstrated in **Figure I**. The following data were extracted: title, lead author, publication year, study purpose, and neighborhood characteristic predictors. Extracted data were collated and stored on an excel spreadsheet coding matrix.

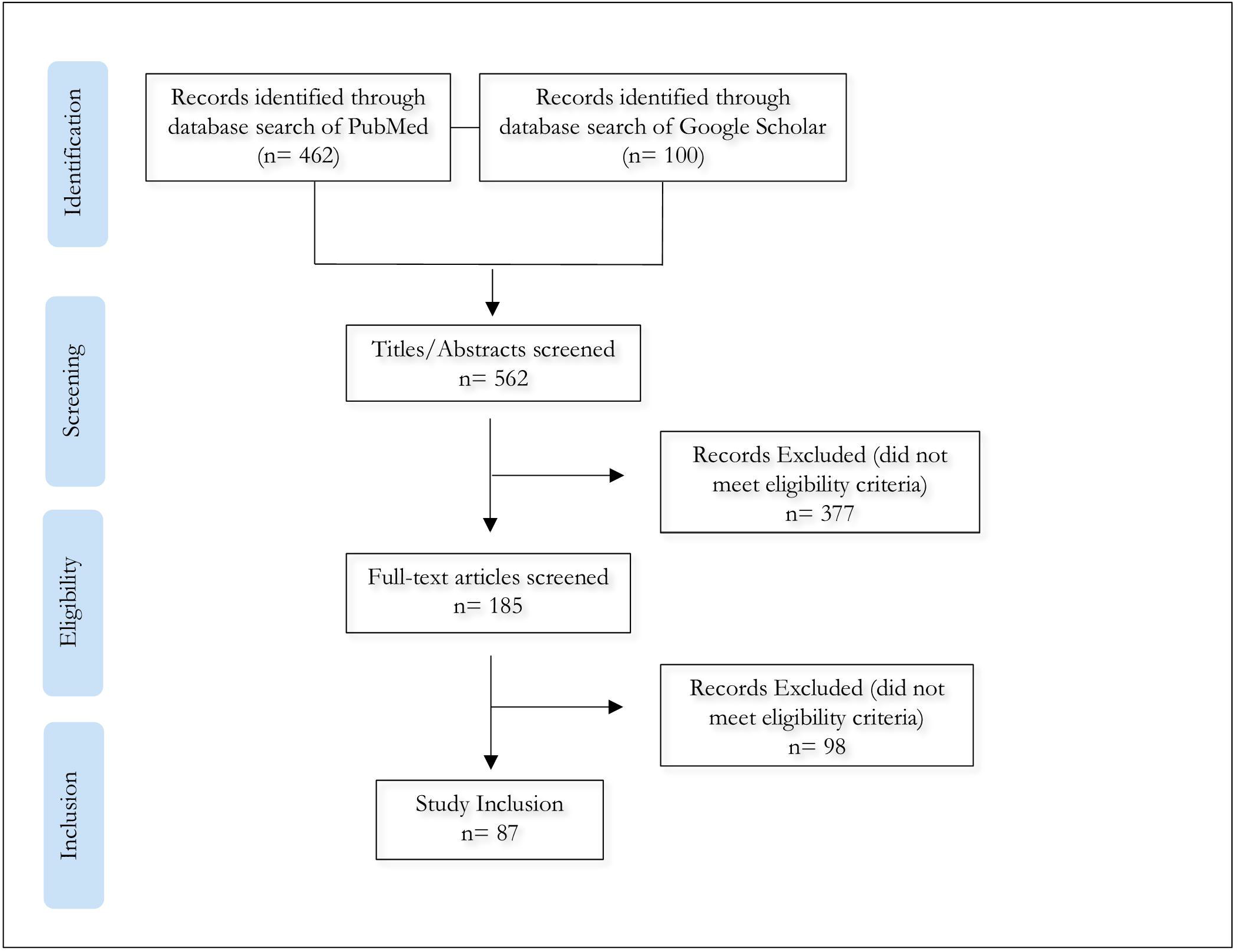

## 3. Results

A total of 87 studies met the inclusion criteria for our review; the oldest of which was published in March 2020, and the most recent in August 2022. A summary table of study characteristics and general information on articles selected for review can be found as supplementary material, labeled “*Additional File 1*”. Overwhelmingly, 100% (87 of 87) of included papers positively reported an influence of neighborhood environment on well-being during the COVID-19 pandemic.

Our research indicates that those living in disadvantaged environments experienced multiple challenges that made them more prone to the severity of COVID-19 infections. Overall, all 87 included studies reported that neighborhood environments influenced health outcomes and population well-being during the COVID-19 pandemic. Moreover, social, biophysical, and built environment exposures were found to be key factors in COVID-19 vaccination success and uptake in 75% of selected studies (65 of 87); 24% (21 of 87) found neighborhood environments had a direct influence on routine childhood vaccination uptake over the pandemic period. Largely, studies argued that developing targeted policies to improve access to healthcare and vaccination services in disadvantaged neighborhoods would be necessary.

Overall, the quality of healthcare, including vaccination success and uptake, was largely impacted by neighborhood indicators such as walkability, housing, transportation, air quality, and poverty levels. We demonstrate a full list of indicators used for the study in **Table I**.

**Table I.** Neighborhood Environment Classification.

| **Built Environment (n=81)** | **Biophysical Environment (n=29)** | **Social Environment (n=83)** |
| --- | --- | --- |
| Transportation (n=50) | Air quality and environmental toxins (n=14) | Safety<br>i. Crime rates (n=22) |
| Walkability (n=11) | Access to Green space (n=16) | Economic stability<br>i. Persistent poverty, household income, employment, housing stability/homelessness (n=68) |
| Housing (n=54) | Climate (n=18) | Politics<br>i. Politicization of COVID-19 related care (n=55) |
| Access to healthcare, insurance coverage (n=72) |  | Mistrust/misinformation<br>i. Governmental mistrust (n=7)<br>ii. Fear (n=36)<br>ii. Expanse of misinformation<br>-Social media's role (n=18) |
| Food Security (n=44) |  |  |

## 3. Discussion

Although the effects of neighborhood conditions are well established as key indicators of population health^15^, the COVID-19 pandemic has disproportionally impacted populations across the US, exacerbating inequalities across multiple facets of the social, biophysical, and built neighborhood environment. While innovative tools were shown to increase testing and vaccinations for some populations, the lack of reliable high-speed internet access, limited access to mobile devices, and low levels of health literacy remained major challenges that hindered access among disadvantaged neighborhoods. A substantial number of individuals faced difficulties in utilizing technology to register for vaccinations, and the distribution of vaccination sites was not equitable in these communities, even though they were experiencing the highest rates of morbidity and mortality from COVID-19. These became significant barriers to achieving vaccine equity. For instance, while policies and protocols were implemented to improve the accessibility of health services and reduce adverse neighborhood influences (i.e., mobile services for testing and vaccination), several studies showed that significant inequalities were reported in terms of access to these services during the pandemic.^16,17^ Moreover, persistent poverty, lack of reliable/affordable housing, poor walkability, and increasing crime rates have contributed to overcrowding and food insecurity, prohibiting low-income families from implementing COVID-19 prevention measures.^18-21^ Many impoverished and marginalized communities lacked social support and as a result, were more susceptible to severe health outcomes due to neglected pandemic protective measures. Initiatives and supports, such as digital literacy programs, might be necessary to empower residents in these disadvantaged neighborhoods to utilize online health services and information.

Public trust in vaccinations, treatment, and prevention solutions plays a key role in establishing a healthy community.^22^ However, during the COVID-19 pandemic, the spread of misinformation, distrust, and vaccine hesitancy posed significant challenges to COVID-19 disease prevention and containment measures.^22^ During the pandemic, examined studies revealed that unaddressed parental fears and concerns contributed to a decrease in overall vaccine uptake which worsened the public’s trust in vaccine safety and effectiveness (i.e., both routine childhood vaccinations and COVID-19 vaccination). This study argues that targeted public health initiatives and community-based policy development to regain public trust and establish reliable sources should be employed to foster vaccination acceptance and adoption of other mitigation measures.

While the biophysical environment is not as frequently addressed in current literature, it is closely interconnected with other domains of the neighborhood environment, making it a crucial area of study. Therefore, policymakers need to give greater consideration to biophysical factors to ensure population well-being. This includes enhancing access to greenspaces, addressing air quality improvement issues, and promoting awareness of climate change. This study suggests that future work should focus on the impacts of these factors and related ones.

Furthermore, addressing long-standing challenges in health outcome disparities for individuals living in disadvantaged neighborhoods requires a comprehensive approach. As many of these challenges are fueled by deep-seated structural and economic inequities, tackling these disparities can be achieved through enhancing the overall health and economic well-being of underserved individuals. In the future, a diverse array of initiatives, both within and outside the healthcare system, will play a crucial role in minimizing disparities among individuals living in disadvantaged and underserved neighborhoods.

Moving forward, it is necessary to consider comprehensive policy alternatives to address the complex challenges posed by neighborhood environments during the COVID-19 pandemic. Notably, there is a need for targeted interventions aimed at bridging the digital divide, ensuring equitable access to technology, and providing digital literacy programs for residents in disadvantaged neighborhoods. Additionally, policymakers should prioritize the fair distribution of healthcare services, overcoming geographical and contextual barriers that impede access for potential users. Various optimization techniques and location-allocation models can contribute to the efficient distribution of resources in this regard.

Looking beyond immediate concerns, long-term strategies should focus on addressing structural inequities by investing in affordable housing, improving walkability, and implementing crime prevention measures. A holistic approach necessitates collaboration between public health agencies, local governments, and community organizations to develop and implement policies that promote population health and well-being. Moreover, establishing public trust requires proactive communication strategies, countering misinformation, and community engagement initiatives. As we chart the course ahead, policymakers, researchers, and community advocates must work collaboratively to incorporate the lessons learned from this pandemic into future policy development, ensuring that the diverse elements of neighborhood environments are thoroughly considered to create resilient, equitable, and healthy communities.

This future call to action emphasizes the importance of an inclusive and forward-thinking approach to policy development, with a focus on addressing the intricate interplay of social, biophysical, and built environment factors to enhance overall community health.

## 4. Conclusions

The COVID-19 pandemic has disproportionally impacted populations across the US, exacerbating present inequalities in all facets of the neighborhood environment. Although the effects of neighborhood conditions are well established as key indicators of population health^15^, the COVID-19 pandemic exacerbated related consequences for deprived communities. As expected, our results suggest that those living in less-than-favorable environments faced a plethora of burdens, making them more prone to the severity of COVID-19 infection.

Our review findings support the argument that various neighborhood environment characteristics substantially impacted population health during the COVID-19 pandemic. Moreover, we propose our findings establish cause for the consideration of the effects of neighborhood environments in intervention strategies for pandemic recovery.

Our results suggest that elements of the built environment strongly affected population health over the pandemic period. Although using mobile services for testing and vaccination aimed to improve the accessibility of health services and reduce adverse neighborhood influences, reviewed studies showed a considerable inequality in terms of access to care during the pandemic. Though these innovative tools were shown to increase testing and vaccination uptake for some populations, reliable high-speed internet access, access to mobile devices, and lack of health literacy remained major challenges preventing their success among disadvantaged neighborhoods. Results showed that the pattern of impacts on population well-being due to infrastructure in built environments is not necessarily the same as in the pre-pandemic era; rather, worsened with added pandemic-related stressors.

Trust in reliable treatment and prevention solutions is key to establishing a healthy community; other efforts will be inconclusive without trust. During the pandemic, examined studies revealed unaddressed parental fears and concerns worsened the public’s trust in vaccine safety and effectiveness (i.e., both routine childhood vaccinations and COVID-19 vaccination), decreasing overall vaccine uptake and subsequent disease protection. We argue that targeted public health initiatives to regain public trust and establish reliable sources should be employed to foster vaccination acceptance and adoption of other COVID-19 mitigation measures.

## Data Availability

All data produced in the present work are contained in the manuscript.

## Author Contributions

BMW conceptualized and supervised the study and drafted, reviewed, and edited the manuscript. WB conceptualized the study, and drafted, reviewed, and edited the manuscript. OO conceptualized the study, and drafted, reviewed, and edited the manuscript. RP drafted, reviewed, and edited the manuscript. FK drafted, reviewed, and edited the manuscript. SH reviewed and edited the manuscript. AS-N drafted, reviewed, and edited the manuscript, supervised the study, and acquired funding.

## Ethical Approval

There is no ethical approval process to disclose, as this research is not considered human subjects research.

## Appendix

### Supplementary Material

**Additional File 1**. Microsoft Word-document .docx, Table S1. Summary Table Representing the Characteristics of Selected Studies

## References

1. World Health Organization. Coronavirus (COVID-19) outbreak [Internet]. World Health Organization; 2022. Available from: https://www.who.int/westernpacific/emergencies/covid-19

2. Johns Hopkins Coronavirus Resource Center. COVID-19 Dashboard [Internet]. 2022. [cited 2023Dec11]. Available from: https://coronavirus.jhu.edu/map.html

3. Dahlberg LL, Krug EG. Violence: a global public health problem. In: Krug E, Dahlberg LL, Mercy JA, Zwi AB, Lozano R, eds. World Report on Violence and Health. Geneva, Switzerland: World Health Organization; 2002:1–21.

4. Kasela, S., Ortega, V.E., Martorella, M. et al. Genetic and non-genetic factors affecting the expression of COVID-19-relevant genes in the large airway epithelium. Genome Med 13, 66 (2021). 10.1186/s13073-021-00866-2

5. Zhang, X., Smith, N., Spear, E., et al. Neighborhood characteristics associated with COVID-19 burden—the modifying effect of age. J Expo Sci Environ Epidemiol 31, 525–537 (2021). 10.1038/s41370-021-00329-1

6. Healthy People 2030. Neighborhood and built environment [Internet]. Neighborhood and Built Environment - Healthy People 2030. [cited 2022Nov21]. Available from: https://health.gov/healthypeople/objectives-and-data/browse-objectives/neighborhood-and-built-environment

7. Brakefield WS, Olusanya OA, Shaban-Nejad A. Association between neighborhood factors and adult obesity in Shelby County, Tennessee: Geospatial Machine Learning Approach. JMIR Public Health and Surveillance. 2022;8(8).

8. DeRouen MC, Tao L, Shariff-Marco S, Yang J, Shvetsov YB, Park S-Y, et al. Neighborhood obesogenic environment and risk of prostate cancer: The multiethnic cohort. Cancer Epidemiology, Biomarkers &amp; Prevention. 2022;31(5):972–81.

9. Stafford M, Marmot M. Neighbourhood deprivation and health: does it affect us all equally? International journal of epidemiology. 2003 Jun 1;32(3):357–66. pmid:12777420

10. Knies G, Melo PC, Zhang M. Neighbourhood Deprivation, Life Satisfaction, and Earnings: Comparative Analyses of Neighbourhood Effects at Bespoke Scales. Urban Studies. 2020 Nov; 10.1177/0042098020956930

11. Piccoli L, Dzankic J, Ruedin D, Jacob-Owens T. Restricting Human Movement During the COVID-19 Pandemic: New Research Avenues in the Study of Mobility, Migration, and Citizenship. Int Migr Rev. 2023;57(2):505–520. doi:10.1177/01979183221118907

12. Zhong X, Zhou Z, Li G, Kwizera MH, Muennig P, Chen Q. Neighborhood disparities in COVID-19 outcomes in New York City over the first two waves of the outbreak. Ann Epidemiol. 2022; 70:45–52. doi: 10.1016/j.annepidem.2022.04.008

13. Carrión, D., Colicino, E., Pedretti, N.F. et al. Neighborhood-level disparities and subway utilization during the COVID-19 pandemic in New York City. Nat Commun 12, 3692 (2021). 10.1038/s41467-021-24088-7

14. Hong B, Bonczak BJ, Gupta A, Thorpe LE, Kontokosta CE. Exposure density and neighborhood disparities in COVID-19 infection risk. Proc Natl Acad Sci USA. 2021;118(13): e2021258118. doi:10.1073/pnas.2021258118

15. Choo J, Kim HJ, Park S. Neighborhood Environments: Links to Health Behaviors and Obesity Status in Vulnerable Children. West J Nurs Res. 2017;39(8):1169–1191. doi:10.1177/0193945916670903

16. Shah GH, Shankar P, Schwind JS, Sittaramane V. The Detrimental Impact of the COVID-19 Crisis on Health Equity and Social Determinants of Health. J Public Health Manag Pract. 2020;26(4):317–319. doi:10.1097/PHH.0000000000001200

17. McGuire AL, Aulisio MP, Davis FD, et al. Ethical Challenges Arising in the COVID-19 Pandemic: An Overview from the Association of Bioethics Program Directors (ABPD) Task Force. Am J Bioeth. 2020;20(7):15–27. doi:10.1080/15265161.2020.1764138

18. Flor LS, Friedman J, Spencer CN, et al. Quantifying the effects of the COVID-19 pandemic on gender equality on health, social, and economic indicators: a comprehensive review of data from March 2020 to September 2021. Lancet. 2022;399(10344):2381–2397. doi:10.1016/S0140-6736(22)00008-3

19. K. C. M, Oral E, Straif-Bourgeois S, Rung AL, Peters ES (2020) The effect of area deprivation on COVID-19 risk in Louisiana. PLoS ONE 15(12): e0243028. 10.1371/journal.pone.0243028

20. Lima FT, Brown NC, Duarte JP. Understanding the Impact of Walkability, Population Density, and Population Size on COVID-19 Spread: A Pilot Study of the Early Contagion in the United States. Entropy (Basel). 2021;23(11):1512. Published 2021 Nov 14. doi:10.3390/e23111512

21. Campedelli et al, Disentangling community-level changes in crime trends during the COVID-19 pandemic in Chicago. https://www.ncbi.nlm.nih.gov/pmc/articles/PMC7590992/

22. Sapienza A, Falcone R. The Role of Trust in COVID-19 Vaccine Acceptance: Considerations from a Systematic Review. Int J Environ Res Public Health. 2022;20(1):665. Published 2022 Dec 30. doi:10.3390/ijerph20010665

