## Additional File 1 for "The Impacts of Neighborhood Disparities on US Population Health During the COVID-19 Pandemic: A Literature Review and Policy Analysis for Future Response"

***Table S1. Summary Table Representing the Characteristics of Selected Studies***

| First Author, Year | Study Purpose | Examined Neighborhood Environment | Author’s Conclusion |
| --- | --- | --- | --- |
| ^1^ Reichert, 2020 | Evaluated the impact of neighborhood environments, particularly built environments, on human mental health in everyday life. | Built, Biophysical, Social | Results suggest that knowledge of urban green spaces, social contacts and physical activity is crucial for both city planning and healthcare as it informs on how to build environments that support healthy living. |
| ^2^ Keenan, 2021 | Examined activities and aspects of the built and biophysical environment associated with climate change and the ongoing COVID responses in the United States. | Built, Biophysical | Findings highlight elements of physical and social parameters of the built environment that may help to inform future disaster, public health, and climate change response. |
| ^3^ Pelling, 2021 | Examined lessons learned from the COVID-19 pandemic for opportunities in enhanced research and action on inclusive urban resilience to climate change. | Built, Biophysical, Social | Results suggest three research priorities: dynamic and compounding vulnerability, systemic risk, and risk root cause analysis. These help to identify affordable and healthy housing, social cohesion, minority and local leadership and multiscale governance for COVID-19 recovery and innovate climate change response and renewal. |
| ^4^ Keenan, 2020 | Provides a perspective through the lens of the built environment on the reciprocal relationships between public and private sector resilience planning activities and the ongoing COVID responses in the United States. | Built, Biophysical | Results suggest that many parameters of the built environment may be useful in informing future disaster, public health, and climate change preparations and responses. |
| ^5^ Landau, 2021 | Uses qualitative data from semi-structured interviews with 34 civic environmental stewardship groups in New York City to explore their role in building adaptive capacity in order to better understand how past crises have impacted stewardship groups' response to COVID-19. | Built, Biophysical, Social | Conclusion suggests that uncovered strategies learned from past public health crises enable stewardship groups to respond to disaster in a way that builds their organizational adaptive capacity, in addition to providing tools for future response and prevention. |
| ^6^ Cheshmehzangi, 2021 | Explored potential development and shifts in the built environment due to COVID-19. | Built, Biophysical, Social | Results may help develop shared practices for public health response, with focus on improving built environments. |
| ^7^ O’Hara, 2021 | Analyzed lessons learned from the COVID-19 pandemic for opportunities in enhanced research and action to address environmental disparities, particularly related to food insecurity. | Built, Biophysical, Social | Conclusion suggests that community-centered strategies may best help shift environmental disparities, such as the food justice discourse from food access to the broader goal of community empowerment. |
| ^8^ Carter-Pokras, 2020 | Discussed the role of epidemiology in the development of immunization policy and successful intervention in public health practice for both routine childhood and COVID-19 vaccinations. | Built, Social | Conclusion suggests that epidemiology remains essential for informing policy and decision making to prevent and respond to vaccine-preventable diseases. Identifying potential determinants of low vaccination coverage (i.e. environmental stressors) is crucial for monitoring the impact of vaccinations on infectious disease incidence and vaccine acceptance by clinicians, parents, and patients. |
| ^9^ Omer, 2021 | Report examined COVID-19 vaccine uptake in the USA, the consequences of low vaccination rates, and recommendations for the improvement of COVID-19 vaccine confidence and uptake. | Built, Social | Conclusion presents a coordinated, evidence-based education, communication and behavioral intervention strategy to improve the success of the United States’ COVID-19 vaccine programs. |
| ^10^ DeSilva, 2022 | Compared trends in pediatric vaccination before and during the pandemic and to evaluate the proportion of children up to date (UTD) with vaccinations by age, race, and ethnicity. | Built, Social | Results suggest that, as of September 2020, childhood vaccination rates were significantly lower than 2019 levels. Conclusions argue for interventions to promote catch-up vaccination, particularly in populations at most vulnerable risk. |
| ^11^ Olusanya, 2021 | Explored and examined barriers to pediatric vaccine uptake behaviors during the COVID-19 pandemic. | Built, Social | Results suggest a multifaceted, multidisciplinary approach involving science, engineering, and social sciences should be incorporated to improve childhood vaccine uptake and comprehend the drivers for vaccine hesitancy, refusal, and delay; including the application of Machine Learning and Artificial Intelligence to (a) identify trends, patterns, and prevalence of childhood vaccine uptake and vaccine-preventable illnesses, (b) investigate psychosocial factors and disparities influencing the receipt of vaccines, and (c) examine the interface between vaccine-preventable disease outbreaks and vaccine hesitancy/refusal. |
| ^12^ Shet, 2022 | Examined the impact of the COVID-19 pandemic on routine immunization. | Built, Social | Results suggest there is evidence to support a sizeable interruption in immunization distribution due to the COVID-19 pandemic. As such, there is urgent need for ongoing assessment of recovery, catch-up vaccination strategy implementation for vulnerable populations, and ensuring vaccine coverage equity and health system resilience. |
| ^13^ Schaffer, 2020 | Discussed and analyzed the social barriers to implantation of a successful COVID-19 immunization program in early 2020. | Social | Conclusion argues the groundwork for public acceptance of a COVID-19 vaccine must be carefully started before a vaccine becomes available. |
| ^14^ Brunson, 2020 | Explores the relationship between social and behavioral research and population COVID-19 vaccination acceptance in early 2020. | Social | Author’s argue for the consideration of social and economic disparities in COVID-19 vaccination program development. |
| ^15^ Weintraub, 2020 | Discussed and described lessons learned from past pandemics and vaccine campaigns about the path to successful vaccine delivery to bolster COVID-19 vaccination rollout efforts in late 2020. | Built, Social | Results suggest lessons of past pandemics indicate governments should invest in evidence-informed strategies to ensure that COVID-19 vaccines protect individuals and minimize disruption to health services and livelihoods. |
| ^16^ Schoch-Spana, 2021 | Examined the major challenges and opportunities associated with early COVID-19 vaccination campaigns to provide empirically-informed recommendations to advance public understanding of, access to, and acceptance of vaccinations. | Built, Social | Conclusion suggests that the development of vaccines is only part of the answer to the COVID-19 pandemic; widespread public acceptance of vaccines is also key. Establishing trust and acceptance is a complex social endeavor that requires the efforts of US policymakers, federal, state, and local public health officials, private funders, professional and community organizations, university researchers, and nontraditional partners. |
| ^17^ Christie, 2021 | Discusses the need for local decision-makers to assess the following factors to inform layered prevention strategies for COVID-19 vaccination coverage: 1) level of SARS-CoV-2 community transmission, 2) health system capacity 3) vaccination coverage, 4) capacity for early detection of increases in COVID-19 cases, and 5) populations at risk for severe outcomes from COVID-19. | Built | Conclusion argues that though increasing COVID-19 vaccination coverage remains the most effective means to control the pandemic, additional layered prevention strategies are needed in the short-term to minimize morbidity and mortality. |
| ^18^ Ellingson, 2022 | Explores and proposes a multi-level framework for understanding the factors that influence provider recommendations at the individual, interpersonal and community level. | Built, Social | Conclusion suggests health care providers play an essential role in vaccination uptake and stand to have meaningful impact to ensuring that adolescents who missed opportunities for routine administration of vaccines during the COVID-19 pandemic are brought up to date. |
| ^19^ Reid, 2021 | Presents observations of the UK, US, and Israel’s experiences in rolling out COVID-19 vaccination programs with mentions of both built and social environment implications. | Built, Social | Results suggest a need for localized public health systems to identify and diminish vaccine hesitancy by building trust and engagement with vulnerable communities. |
| ^20^ Alfieri, 2021 | Compared and analyzed hesitancy toward COVID-19 vaccines for children of various sociodemographic groups in major metro areas. Additionally, highlighted the need to understand how parents obtain information about COVID-19. | Built, Social | Results suggest the highest rates of hesitancy toward COVID-19 vaccination programs were found in demographic groups most severely affected by the pandemic. These groups require targeted outreach efforts from trusted sources to promote equitable uptake of COVID-19 vaccines. |
| ^21^ He, 2021 | A cross-sectional mobile phone-based survey at Children’s Hospital Los Angeles querying parents on perspectives related to vaccines before and during the pandemic. The study aimed to understand the impact of the COVID-19 pandemic on routine childhood vaccine hesitancy, COVID-19 vaccine hesitancy, and key contributing demographic factors. | Built, Social | Results found that routine childhood vaccine hesitancy increased during the COVID-19 pandemic, mainly due to increased risk perception. Key contributing factors behind both routine and COVID-19 vaccine hesitancy included household income and race. Ultimately, understanding factors behind routine childhood vaccine hesitancy is crucial to maintaining pediatric vaccination rates and confidence during/after the COVID-19 pandemic. |
| ^22^ Fisher, 2022 | Examined 1) the extent of which racial/ethnic differences both jointly and independently predict pediatric COVID-19 parental vaccine hesitancy, and 2) assessed similarities and differences in the prominence of these predictors for Hispanic and non-Hispanic Asian, Black, and White parent populations. | Built, Social | Findings highlight the importance of multipronged population targeted approaches to increase pediatric COVID-19 vaccine uptake. Conclusions argue for these measures to include integrating health science literacy with safety/efficacy messaging, tailored communication efforts for parents who express hesitancy, and interventions developed in partnership with trusted community organizations. |
| ^23^ David, 2022 | A retrospective assessment of the relationship between ethnicity, social determinants of health, and measures of pediatric health outcomes during the COVID-19 pandemic. | Built, Social | Results suggest that the COVID-19 pandemic disproportionately affected patients with unmet social determinants in obtaining routine pediatric care including immunizations. |
| ^24^ Finney Rutten, 2020 | Reviewed, summarized, and encouraged use of interpersonal, individual-level, and organizational interventions within clinical organizations to address evident lack of confidence in COVID-19 vaccination to improve population uptake. | Built, Social | Conclusion calls for the use of evidence-based strategies at the  organizational, interpersonal, and individual levels in clinical organizations to increase uptake of COVID-19 vaccination. |
| ^25^ Khubchandani, 2021 | Conducted a comprehensive and systematic national assessment of COVID-19 vaccine hesitancy in the United States. | Built, Social | Findings suggest that, before COVID-19 vaccination rollout, 22% of study respondents were hesitant to receive the vaccines if/when they were made available. Interestingly, differences in vaccine hesitancy were based on sociodemographic characteristics including sex, race, ethnicity, education, income, employment status, place of residence, political affiliation, and personal perceived risk of infection. As such, authors argue that evidenced-based educational and policy-level interventions that are necessary to promote COVID-19 immunization programs. |
| ^26^ Wiysonge, 2022 | Evaluated whether lessons learned from past public health emergencies could aid in combatting COVID-19 vaccination hesitancy. | Built, Social | Conclusion suggests that vaccine hesitancy is a complex social process involving multiple webs of influence, meaning, and logic. Population view of vaccination is usually a dynamic personal process, unfolding and changing overtime. As such, public health professionals should continually investigate determinants of vaccine hesitancy to develop targeted hesitancy mitigation strategies. |
| ^27^ Shah, 2020 | Discussed the impacts of the COVID-19 pandemic on health equity and social determinants of health. | Built, Social | Results and conclusions suggest the impacts of COVID-19 infection are significantly higher for most vulnerable communities (i.e., elderly, impoverished, those with barriers to healthcare access, chronically ill, racial/ethnic minorities, etc.). |
| ^28^ Anderson, 2022 | Discussed reasons behind low COVID-19 vaccine acceptance in pregnant and postpartum individuals in vulnerable communities. | Built, Social | Conclusion argues for the continued development of a robust and effective healthcare system that provides equitable care to all perinatal patients regardless of demographics. Authors argue this is essential to understanding how trust and other psychological determinants of COVID-19 vaccine hesitancy shapes individuals' vaccination beliefs behavior. Authors further argue the importance of early behavioral intervention for pregnant individuals, as hesitant beliefs may impact future vaccination decisions for their children. |
| ^29^ Kiefer, 2022 | A cross-sectional study to assess the frequency and associated characteristics of COVID-19 vaccine hesitancy among pregnant and postpartum individuals. | Built | Findings suggest COVID-19 vaccine hesitancy was frequent among pregnant and postpartum individuals. Interestingly, findings also uncovered those who face barriers to accessing healthcare were more likely to be vaccine hesitant. Moving forward, these results can inform interventions to increase COVID-19 vaccine uptake in pregnant individuals, particularly for those in vulnerable communities. |
| ^30^ McGuire, 2020 | Discussed resource allocation and related ethical challenges to healthcare access including, 1) how to define benefit, 2) how to handle informed consent, 3) the special needs of pediatric patients, 4) how to engage communities in these difficult decisions, and 5) how to mitigate concerns of discrimination and the effects of structural inequities. | Built, Social | Conclusion suggests bioethicists and healthcare professionals should continue to learn the lessons of the COVID-19 pandemic, including consideration of various environmental stressors, and rapidly put them into practice to be better prepared for the next public health emergency. |
| ^31^ Garg, 2021 | Reviewed current literature on the impact of the COVID-19 pandemic on the LGBTQ+ population on the basis of social determinants to build recommendations to address vaccine hesitancy. | Built, Social | Conclusions find the LGBTQ+ population has suffered from increased systemic discrimination, oppression, and structural health inequities through the COVID-19 pandemic. There is urgent need for a collaborative effort between federal governments, healthcare policymakers, healthcare providers, and mass media providers to build trust and provide accurate information on vaccine safety and efficacy to protect this vulnerable group. |
| ^32^ Newman, 2022 | Provides a comprehensive review of determinants of COVID-19 vaccine hesitancy among marginalized populations in the United States and Canada. | Built, Social | Conclusions argue findings contribute to ongoing efforts advancing the conceptualization of COVID-19 vaccine hesitancy, identify evidence gaps, and support recommendations for future research and practice. |
| ^33^ Oberg, 2022 | Examined the pediatric impacts of the COVID-19 pandemic in the United States by highlighting systemic/institutional inequities accentuated by the pandemic for vulnerable populations, in addition to evaluating disruptions in health care access, childcare, and education. | Built, Social | Findings argue that immediate and long-term effects of the COVID-19 pandemic on children and young people should not be underestimated, as the crisis has amplified inequities, increased stress and strain on families and existing resources. |
| ^34^ Salmon, 2021 | Reflected upon equity challenges hindering COVID-19 vaccination rollout. | Built, Social | Conclusion argues there is a need for multifaceted communication plan that is responsive to the concerns and values of all groups, implemented by federal, state, and local governments. Moreover, healthcare providers are well positioned to effectively disseminate trusted information to patients for informed decision-making. Ultimately, an equitable COVID-19 immunization program could help reduce the risks associated with the COVID-19 pandemic for vulnerable populations. |
| ^35^ Vicetti Miguel, 2022 | Reviews the relationship between COVID-19 and pediatrics health disparities, access to healthcare, pharmaceutical interventions, and clinical research in the United States. | Built, Social | Conclusion presents potential strategies to communicate scientific data in ways that do not promote racism or biological susceptibility themes, while addressing pediatric disparities in clinical infectious disease research. |
| ^36^ Cholera, 2020 | Evaluates the impact of COVID-19 on children of immigrant families to highlight opportunities for advocacy/action for healthcare providers, hospitals and healthcare systems, and policymakers to mitigate risks for this vulnerable group. | Built, Social | Findings suggest COVID-19 mitigation strategies, including vaccination programs, must incorporate policy and public health approaches that are compassionate, culturally relevant, and equitable for all, especially for children in immigrant families. |
| ^37^ Flor, 2022 | Explored indirect effects of COVID-19 on gender disparities. | Built, Social | Results find significant gaps in widespread inequalities between women and men during the COVID-19 pandemic. |
| ^38^ Abrams, 2020 | Examined the impacts of social determinants of health - including poverty, physical environment (i.e., smoke exposure, homelessness), and race or ethnicity - on COVID-19 outcome. | Built, Social | Conclusion argues for consideration of social determinants of health as part of pandemic research priorities, public health goals, and policy implementation. |
| ^39^ Jones, 2021 | Investigates the United States’ capacity to contain disease outbreaks. Additionally, outlines the need for public health professional planning to achieve greater health equity for future outbreaks by addressing underlying economic and social conditions to provide meaningful access to preventive care for all. | Built, Biophysical, Social | Conclusion suggests officials are at a critical moment of opportunity to take meaningful action to remedy social inequities in future public health emergencies based on lessons learned from the COVID-19 pandemic. |
| ^40^ Fronteira, 2021 | Evaluated the syndemic theory that translates the cumulative and intertwined factors between different epidemics to argue that COVID-19 pandemic is a health issue of a syndemic nature. Moreover, argues the failure to acknowledge this characteristic nature of the pandemic has contributed to weakened policy-making and public health response, allowing for accumulation of social disadvantages. | Built, Biophysical, Social | Conclusions suggest acknowledgement of the syndemic nature of the COVID-19 pandemic in the development of mitigation strategies would allow public health professionals to combat threats to livelihood including economic strain, food insecurity, mistrust/misinformation, etc.. |
| ^41^ Rodrigues, 2022 | Explores insight from previous literature to determine what works to increase vaccine uptake and how best practices can be applied to increase COVID-19 vaccine uptake. | Built, Social | Findings suggest the various reasons for vaccine hesitancy are quite complex. As such, a multifaceted approach is needed to address hesitancy. Ultimately, understanding the factors that affect vaccine hesitancy could aid to address widespread hesitancy and contribute to an increase in vaccine uptake. |
| ^42^ Kumar, 2022 | Examined the wellbeing of pediatric populations in urban and rural areas during the COVID-19 pandemic. | Built, Social | Results exhibit persistent unmet needs in urban and rural areas during the COVID-19 pandemic, with higher trends of health and social needs in rural areas even in light of higher levels of social service use. |
| ^43^ Parimi, 2022 | Compared vaccine hesitancy prior to the emergence of COVID-19 against recent reports on vaccine hesitancy related to COVID-19 vaccines. | Social | Conclusion suggests a better understanding of the social reasons driving vaccine hesitancy will hopefully inform strategies to promote vaccine safety and increase the number of vaccinated individuals. |
| ^44^ Afolabi, 2021 | Examined and described the role of community participation/partnership is useful to promote COVID-19 vaccination acceptance. | Social | Findings suggest the engagement of community stakeholders such as community-based organizations, leaders religious groups, and media teams could be extremely valuable to address COVID-19 hesitancy and improve vaccine acceptance. Moreover, conclusions argue community members should be actively involved in the design and implementation of COVID-19 vaccine distribution plans; such involvement stands to empower populations from within and improve vaccine hesitancy overall. |
| ^45^ Zhang, 2021 | Explored and summarized structural and attitudinal barriers to vaccine acceptance in the United States. | Built, Social | Conclusions suggest that, while the logistics of distributing a two-dose vaccine are challenging, all vaccines must be distributed fairly to achieve equal protection and reduce disparities. Furthermore, more resources are needed to address attitudinal barriers, as these types of hurdles are more difficult to overcome compared to structural. |
| ^46^ Davenhall, 2022 | Examines and explores the role of Geographic Information Systems in response to the COVID-19 pandemic and various related social determinants, particularly for case identification and vaccination distribution. | Built, Social | Conclusion highlight potential COVID-19 response advancements made possible by Geographic Information Systems. |
| ^47^ Afzal, 2022 | A survey-based study to understand the factors affecting COVID-19 vaccine attitudes among health care professionals in NYC Health and Hospitals. | Built, Social | Findings suggest that local social influences likely have a substantial impact on COVID-19 vaccine uptake among health care professionals in NYC Health and Hospitals. As such, local and demographic factors should be strongly considered when preparing vaccination promotion messages or campaigns. |
| ^48^ El-Mohandes, 2021 | Assesses attitudes towards COVID-19 vaccination among adults in the four largest metropolitan areas in the United States. | Built, Biophysical, Social | Findings provide an understanding of perceptions underlying COVID-19 vaccine acceptance in the United States, uncovered with the utilization of a novel COVID-19 vaccine acceptance scale – ‘COVID-VAC’ – while describing motivational factors that could be used to sway those who are hesitant. |
| ^49^ Tharpe, 2021 | Identifies barriers to vaccine uptake and describes harm reduction measures designed to improve uptake of vaccines. | Built, Social | Conclusions argue that healthcare providers are expected to be knowledgeable about vaccine benefits, recommendations, side effects, and potential adverse reaction and communicate these efficiently to all patients. Moreover, socioeconomic factors, social policies, and historic experiences related to racism are identified as barriers to vaccine acceptance and should be considered in future policy development and decision-making. |
| ^50^ Bloom, 2021 | Discusses the importance of adopting a broad societal perspective as an appropriate standard for health technology assessment and the potential role of COVID-19 in advancing this perspective. | Social | Findings suggest public acceptance of COVID-19 vaccines largely depends on the transparency and trustworthiness of political leaders charged with overseeing the government’s role in vaccine research and development, manufacturing, and delivery. |
| ^51^ Isasi, 2021 | Examines the impact/implications of the COVID-19 pandemic on patients, families, and communities. | Built, Biophysical, Social | Results support that patients, families, and communities have been further impacted by the multisectoral responses to the COVID-19 pandemic – both positively and negatively. In particular, many aspects of the pandemic have exacerbated community level problems (i.e. poverty, access to healthcare, etc.) for most vulnerable populations. As such, there is a need for both short- and long-term changes to the United States’ public health response system. Author’s argue that centering the needs of patients, families, and communities must be at the core of any proposed changes. |
| ^52^ Hu, 2021 | Investigated the relationships between the built and social environments and their impact on population health during the COVID-19 pandemic. | Built, Biophysical, Social | Findings suggest the collective impact of environmental determinants increased the risk of COVID-19, particularly for marginalized communities. Author’s argue that these findings can assist public health policymakers and stakeholders in efforts to control and mitigate the effects of the COVID-19 pandemic. |
| ^53^ Wakeel, 2021 | Applied the Weathering Framework to conceptualize COVID-19 as a stressor inflicting profound health implications for African American communities. | Built, Social | Conclusions recommend further population health research, interventions and policies aimed at reducing COVID-19 morbidity and mortality, as well as support for mitigation of the long-term impacts of the pandemic on marginalized communities. |
| ^54^ Firebaugh, 2020 | Explores the vulnerability of the rural Yakima County, Washington region during the COVID-19 pandemic – including an examination of relevant environmental stressors. | Built, Biophysical, Social | Conclusion calls for environmental, structural, social, and policy level responses to support rural communities during public health emergencies such as the COVID-19 pandemic. Multifaceted approaches are key to protecting these regions and must be considered in strategy development. |
| ^55^ Rifat, 2022 | Explores spatial distributions and patterns of COVID-19 cases, mortality rates, and related disparities between urban and rural counties in the contiguous United States. | Built, Biophysical, Social | Findings suggest relationships between social vulnerability components and case/mortality rates vary; however, counties with increased racial and ethnic minorities, higher percentages of minors, and lower median household income were associated with higher COVID-19 case rates and number of deaths. |
| ^56^ Mah, 2021 | Examines the possibility that disparities in social, economic status, and physical environment may have contributed to high mortality rates during the COVID-19 pandemic. | Built, Biophysical, Social | Conclusions recommend a multifaceted response based on county-level data in coordination between health systems and local governments to identify social disparity ‘hotspots’ and using the findings for targeted resource/intervention allocation. |
| ^57^ Magnan, 2021 | Provides a framework to implement relevant strategies, processes, and policies for community and individual approaches to address social determinants of health, social risk factors, and health-related social needs in light of the disparate impact of COVID-19 on vulnerable communities. | Built, Biophysical, Social | Findings suggest community partners and health care organizations should use the Plan-Do-Study-Act (PDSA) cycle to determine what community needs (i.e. social risk factors, environmental stressors, etc.) are of most importance to allow for time/need targeted responses. |
| ^58^ Bogan, 2022 | Highlights impacts of the COVID-19 pandemic on Black children at the individual, family, and school levels. | Built, Social | Conclusions make research, practice, and policy recommendations to journal editors, funding agencies, grant review panels, and researchers on how COVID-19 research should be framed to inform intervention strategies for improving situation of Black children and families. |
| ^59^ Cole, 2022 | Explores the intensified health inequities exposed by the COVID-19 pandemic, and how social determinants caused disproportionately higher rates of infection, morbidity, and mortality in marginalized and disadvantaged communities. | Built, Social | Results find that many barriers must be considered in providing equitable access to COVID-19 care for patients and communities: location and transportation barriers, communication barriers, and hesitation and mistrust regarding uncertainties of a new vaccine to name a few. Moreover, lessons learned from COVID-19 response should be well documented and applied for future emergencies. |
| ^60^ Gil, 2021 | Contextualized COVID-19 inequities through the history of the HIV/AIDS crisis; described LGBTQ+ structural oppression exacerbated by the pandemic; and provided recommendations for medical professionals/institutions to reduce health inequities. | Built, Social | Conclusion suggests that it is crucial to note that social determinants (i.e. underemployment, poverty, etc.) have been found to be connected to greater impacts of COVID-19. Authors argue that individuals, providers, and organizations must fight for equitable care and amplify patients’ voices/experiences, to provide efficient care to most vulnerable populations. |
| ^61^ Brakefield, 2022 | Described the design/development of the Urban Population Health Observatory, an explainable knowledge-based multimodal big data analytics platform. The Urban Population Health Observatory is designed to integrate multimodal big data including social determinants of health, observation of daily living, and population health data consistently across jurisdictions to estimate the incidence/prevalence of health conditions and related risk factors. | Built, Biophysical, Social | Conclusion suggests the development of a multimodal scalable surveillance system to monitor and detect trends, identify social determinants, and deliver rapid early warnings would assist health officials, providers, and researchers in responding and mitigating a public health crisis such as COVID-19. |
| ^62^ Loukaitou-Sideris, 2021 | Sought to understand the scale of homelessness on transit and how transit agencies are responding to the problem, as COVID-19 has exacerbated the rate of homelessness in the United States. | Built, Biophysical, Social | Findings suggest that transit serves as shelter for a high number of unsheltered/homeless individuals, who are more likely than their unhoused peers elsewhere to be structurally disadvantaged. Moreover, there is a need for fostering external partnerships, keeping law enforcement distinct from routine homeless outreach, educating the public, and training transit staff to aid in equitable treatment of unhoused communities facing multifaceted environmental disparity. |
| ^63^ Perrigo, 2022 | Explored minority and low-SES families’ experiences during the COVID-19 pandemic crisis. | Built, Social | Findings suggest the more social resources located in a neighborhood, the more confidence communities have in overcoming a crises, such as the COVID-19 pandemic – even for those classified as low-SES. Authors argue there is a need for further examination of long-term trends to inform meaningful policies to support how low-SES families buffer against COVID-19-related negative impacts and mitigate other inequities/disparities. |
| ^64^ Harrison, 2021 | Examines differences in the effects of COVID-19 on minority communities – with particular focus on the accompanying social and economic impacts of the pandemic. | Built, Social | Conclusion suggests important differences exist in the way various minority groups/neighborhoods experience the progression of COVID-19 infection and outcome. Authors argue future work is needed to fully explore causality of differences, particularly as they relate to most vulnerable communities. |
| ^65^ Chumpa, 2021 | Examines and explores social determinants of COVID-19 related sickness/suffering in the Bronx, New York City based on neighborhood zip codes. | Built, Biophysical, Social | Findings suggest factors ranging from environmental pollution to low access to nutritious foods created a barrier to healthy living for those in vulnerable neighborhoods, leading to increase disease susceptibility. Moreover, language barriers, stigmatization of those diagnosed with COVID-19, and culturally insensitive healthcare delivery worsened the gap for those at highest risk, contributing to greater disease effect. |
| ^66^ Bonotti, 2021 | Provides an overview of the human, economic, social, and political costs of the COVID-19 pandemic. | Built, Biophysical, Social | Conclusion suggests the COVID-19 pandemic has created immense strain on daily activities, including the way people interact with one another. Access to healthcare, transportation changes, housing disparities, food security, air pollution, access to green space, crime, poverty, politics, and mistrust/misinformation are all listed as related to pandemic challenges. |
| ^67^ Chen, 2020 | Explored and modeled spatial factors and determinants of COVID-19 in New York City. | Built, Biophysical, Social | Conclusion suggests policymakers should implement unique policies, preventions measures, and reopening strategies based on localized situations/neighborhoods/regions to mitigate ongoing COVID-19 outbreaks. |
| ^68^ Byttebier, 2022 | Explores various environmental inequalities in light of the COVID-19 pandemic. | Built, Biophysical, Social | Findings present and discuss the characteristics of the determinants which have contributed to the severity of COVID-19 in the United States. |
| ^69^ Barzilay, 2020 | Explored elements of self-reliance, emotion-regulation, interpersonal-relationship patterns and neighborhood-environment during the acute COVID-19 outbreak. | Social | Findings provide evidence that communities living under higher levels of stress (i.e. those living in deprived neighborhood environments) expressed more concerns related to the wellbeing of those around them. |
| ^70^ Bavel, 2020 | Discussed evidence relevant to pandemic management in early 2020, including navigating threats, social and cultural influences on behavior, communication, moral decision-making, leadership, and stress/coping mechanisms. | Built, Biophysical, Social | Conclusions identified insight for effective response to the COVID-19 pandemic in early 2020 by highlighting important gaps in need of filling; including consideration of built, biophysical, and social environments affecting disease transmission. |
| ^71^ Carrión, 2021 | Investigated the association between neighborhood social disadvantages and COVID-19 infections and mortality in the Spring of 2020. | Built, Biophysical, Social | Results display a positive association between neighborhood social disadvantage and number of COVID-19 infections. Findings contribute to the argument that greater impacts of COVID-19 may be strongly correlated with measures of neighborhood structural disadvantage. |
| ^72^ K.C., 2020 | Examined the relationship between neighborhood deprivation and COVID-19 in Louisiana. | Built, Social | Findings reflect that individuals residing in the most deprived neighborhoods had a ~40% higher risk of COVID-19 infection compared to those residing in the least deprived neighborhoods. Authors argue findings should be utilized to promote public health response measures in most vulnerable neighborhoods. |
| ^73^ Anderson, 2022 | Evaluated racial-ethnic residential clustering and it relates to early COVID-19 vaccine rollout. | Built, Biophysical, Social | Results suggest that neighborhood disparities in service provision are key to understanding racial-ethnic inequality in public health emergencies such as the COVID-19 pandemic. |
| ^74^ Benninger, 2021 | Examined how social determinants contribute to neighborhood health and the wellbeing of children who live within them. | Built, Biophysical, Social | Findings present four thematic neighborhood characteristics connected to youth health and well-being. These include: 1) Crime and safety, 2) Housing and the built environment, 3) Social Influence, and 4) Community Activities. Conclusion argues for the consideration of these four thematic categories for future research, policy, and intervention programming. |
| ^75^ Bilal, 2022 | Explored the association between neighborhood-level social vulnerability and COVID-19 vaccination coverage in 16 cities in the United States from December 2020 to September 2021. | Built, Social | Conclusions report vulnerability index domains of socioeconomic status, household composition, and disability showed the strongest associations with vaccination coverage. Ultimately, neighborhood inequities in COVID-19 vaccination distribution and uptake significantly effect health equity and could lead to even wider disparity in other outcomes. |
| ^76^ Choi, 2022 | Evaluated community health workers perspectives on an COVID-19 vaccine outreach program targeted at a homeless population in Los Angeles. | Built, Social | Conclusions suggest health workers/providers are in a unique position to empower unhoused populations to both receive the COVID-19 vaccination and encourage their peers to do the same, in addition to self-empowerment for advocation for long-term employment and housing needs. |
| ^77^ Crist, 2022 | Sought to understand how transportation has impacted health and equity outcomes during the COVID-19 pandemic. | Built, Biophysical, Social | Conclusion suggests the need for further research on how transit usage impacts population health; thus, authors propose a ‘TROLLEY’ study, aimed at informing land use, transportation and health investments, and workplace interventions to bolster population health and wellbeing not only in response to the COVID-19 pandemic, but as a consistent public health priority. |
| ^78^ DiRago, 2022 | Investigated how strongly COVID-19 vaccination levels were associated with socioeconomic and racial/ethnic disparities. | Built, Biophysical, Social | Findings suggest that vaccine rollouts contributed to cumulative disadvantage across the largest cities in the United States. Moreover, populations that were left most vulnerable to COVID-19 benefited least from early expansions in vaccine availability. |
| ^79^ Ezell, 2021 | Discussed how COVID-19 has both inherited and augmented patterns of spatial inequality. | Built, Biophysical, Social | Conclusion outlines relevant steps that should be taken to prevent and reduce spatial inequalities generated by COVID-19. |
| ^80^ Hillis, 2021 | Reports overall COVID-19–associated orphanhood and death of grandparent caregivers for the United States disaggregated by race and ethnicity. | Built, Social | Results suggest high rates of orphanhood, recorded disparities, and state-specific differences show the overlooked burden among vulnerable children throughout the COVID-19 pandemic. |
| ^81^ Mollalo, 2021 | Compiled a database of the percentage of fully COVID-19 vaccinated Americans at the county scale as of 29 July 2021, along with social vulnerability index data as potential significant covariates to determining vaccine uptake. | Built, Social | Conclusion suggests findings should serve as a geospatial reference to support public health decision-makers in forming region-specific policies to monitor COVID-19 vaccination programs. |
| ^82^ Morgan, 2022 | Examined whether demographic factors such as race/ethnicity, perceived discrimination, and medical mistrust were predictors of COVID-19 vaccine hesitancy; and evaluated whether medical mistrust and perceived discrimination potentially mediate the relationship between race/ethnicity and vaccine acceptance/behavior. | Built, Social | Findings indicate the need for public health efforts to address public medical mistrust and experiences of perceived discrimination to combat COVID-19 vaccine hesitancy, particularly within most vulnerable populations. |
| ^83^ Ransome, 2021 | Evaluated whether neighborhood social cohesion is associated with inequalities in COVID-19 diagnosis rate and the extent the association varies across neighborhood racial composition. | Built, Social | Results suggest that neighborhood social cohesion is associated with both higher and lower COVID-19 diagnosis rates. Moreover, the extent of associations varies across Black neighborhood racial composition. Conclusion recommends strategies for reducing inequalities based on the social cohesion and public health intervention framework. |
| ^84^ Sacarny, 2021 | Used neighborhood-level data to estimate inequities in COVID-19 vaccination rates. | Built, Social | Results suggest COVID-19 vaccination rates were disproportionately high in communities with lower burdens of this disease in the 9 largest cities in the United States. Conclusion calls for the opportunity/need for cities across the United States to address vaccination inequities in marginalized communities to ensure widespread population protection. |
| ^85^ Xiao, 2022 | Sought to answer two questions: 1) How does COVID-19 influence the light rail ridership?, 2) Which light rail stations are more vulnerable to COVID-19 and recover faster?. Focused on the vulnerability and resilience of public transit ridership, and the decline of transit ridership during the COVID-19 pandemic to measure vulnerability and the recovery of transit ridership. | Built, Social | Results find that commuting behavior of minoritiy populations is less likely to be influenced by the COVID-19 pandemic, implying that minorities are more exposed to the disease. Furthermore, authors conclude this may be explained in that that the minorities are less likely to be able to “work from home”, and taking public transit may make them more likely to be exposed to COVID-19. |
| ^86^ Zeng, 2022 | An ecological study that sought to examine the effects of population mobility and proportion of older adults on COVID-19 incidence and test whether the interaction between these factors was significant in predicting the spread of COVID-19 in the Deep South of the United States. | Built, Social | Conclusions suggest that population mobility contributed to the geospatial disparities observed in county-level COVID-19 incidence in the Deep South. Among populations with a high percentage of elderly individuals, population mobility has a stronger impact on COVID-19 outbreak than populations of younger average age. As such, authors argue that policies regarding social distancing and travel restrictions should be tailored to vulnerable communities for mitigation of future public health emergencies. |
| ^87^ Gin, 2022 | Explored experiences of Homeless Patient Aligned Care Teams in navigating the tasks of keeping Veterans safe and providing ongoing care from the start of the COVID-19 pandemic to 2021. | Built, Social | Conclusion suggests there is an urgent need for enhanced pandemic preparedness planning, funding for personal protective equipment,  technology to facilitate Veterans’ telehealth access, and strategies to prevent provider burnout. Authors argue these tools are critical to sustaining homeless providers’ capabilities and enhancing response to future public health emergencies. |

^1^ Reichert M, Braun U, Lautenbach S, Zipf A, Ebner-Priemer U, Tost H, et al. Studying the impact of built environments on human

mental health in everyday life: Methodological Developments, state-of-the-art and technological frontiers. Current Opinion in

Psychology. 2020Apr;32:158–64.

^2^ Keenan, J.M. (2021). COVID and Climate: Exploring Categorical Resilience in the Built Environment. In: Linkov, I., Keenan, J.M.,

Trump, B.D. (eds) COVID-19: Systemic Risk and Resilience. Risk, Systems and Decisions. Springer, Cham.

https://doi.org/10.1007/978-3-030-71587-8_15

^3^ Pelling M, Chow WT, Chu E, Dawson R, Dodman D, Fraser A, et al. A climate resilience research renewal agenda: Learning lessons from the covid-19 pandemic for urban climate resilience. Climate and Development. 2021Sep22;14(7):617–24.

^4^ Keenan, J.M. COVID, resilience, and the built environment. Environ Syst Decis 40, 216–221 (2020). https://doi.org/10.1007/s10669-

020-09773-0

^5^ Landau LF, Campbell LK, Svendsen ES, Johnson ML. Building adaptive capacity through civic environmental stewardship: Responding to COVID-19 alongside compounding and concurrent crises. Frontiers in Sustainable Cities. 2021Nov11;3.

^6^ Cheshmehzangi A. Revisiting the built environment: 10 potential development changes and paradigm shifts due to

COVID-19. *J Urban Manag*. (2021) 10:166–75. 10.1016/j.jum.2021.01.002

^7^ O'Hara S, Toussaint EC. Food access in crisis: Food security and covid-19. Ecological Economics. 2021Feb;180:106859.

^8^ Carter-Pokras O, Hutchins S, Gaudino JA, et al. The role of epidemiology in informing United States childhood immunization policy

and practice. Ann Epidemiol. 2021;62:100-114. doi:10.1016/j.annepidem.2020.09.017.

^9^ Omer SB, Benjamin RM, Brewer NT, et al. Promoting COVID-19 vaccine acceptance: recommendations from the Lancet

Commission on Vaccine Refusal, Acceptance, and Demand in the USA. Lancet. 2021;398(10317):2186-2192.

doi:10.1016/S0140-6736(21)02507-1

^10^ DeSilva MB, Haapala J, Vazquez-Benitez G, et al. Association of the COVID-19 Pandemic With Routine Childhood Vaccination

Rates and Proportion Up to Date With Vaccinations Across 8 US Health Systems in the Vaccine Safety Datalink. JAMA

Pediatr. 2022;176(1):68–77. doi:10.1001/jamapediatrics.2021.4251

^11^ Olusanya OA, Bednarczyk RA, Davis RL and Shaban-Nejad A (2021) Addressing Parental Vaccine Hesitancy and Other Barriers to

Childhood/Adolescent Vaccination Uptake During the Coronavirus (COVID-19) Pandemic. Front. Immunol. 12:663074. doi:

10.3389/fimmu.2021.663074

^12^ Shet A, Carr K, Danovaro-Holliday MC, Sodha SV, Prosperi C, Wunderlich J, et al. Impact of the SARS-COV-2 pandemic on

routine immunization services: Evidence of disruption and recovery from 170 countries and Territories. The Lancet Global

Health. 2022Dec21;10(2).

^13^ Schaffer DeRoo S, Pudalov NJ, Fu LY. Planning for a COVID-19 Vaccination Program. JAMA. 2020;323(24):2458–2459.

doi:10.1001/jama.2020.8711

^14^ Brunson EK, Schoch-Spana M. A Social and Behavioral Research Agenda to Facilitate COVID-19 Vaccine Uptake in the United

States. Health Secur. 2020;18(4):338-344. doi:10.1089/hs.2020.0106

^15^ Weintraub RL, Subramanian L, Karlage A, Ahmad I, Rosenberg J. Covid-19 vaccine to vaccination: Why leaders must invest in

delivery strategies now. Health Affairs. 2020Nov19;40(1):33–41.

^16^ Schoch-Spana M, Brunson EK, Long R, et al. The public's role in COVID-19 vaccination: Human-centered recommendations to enhance pandemic vaccine awareness, access, and acceptance in the United States. Vaccine. 2021;39(40):6004-6012. doi:10.1016/j.vaccine.2020.10.059

^17^ Christie A, Brooks JT, Hicks LA, et al. Guidance for Implementing COVID-19 Prevention Strategies in the Context of Varying Community Transmission Levels and Vaccination Coverage. MMWR Morb Mortal Wkly Rep. 2021;70(30):1044-1047. Published 2021 Jul 27. doi:10.15585/mmwr.mm7030e2

^18^ Ellingson, M.K., Bednarczyk, R.A., O’Leary, S.T. et al. Understanding the Factors Influencing Health Care Provider Recommendations about Adolescent Vaccines: A Proposed Framework. J Behav Med (2022). https://doi.org/10.1007/s10865-022-00296-4

^19^ Reid JA, Mabhala MA. Ethnic and minority group differences in engagement with COVID-19 vaccination programs - at Pandemic Pace; when vaccine confidence in mass rollout meets local vaccine hesitancy [published correction appears in Isr J Health Policy Res. 2021 Oct 28;10(1):60]. Isr J Health Policy Res. 2021;10(1):33. Published 2021 May 27. doi:10.1186/s13584-021-00467-9

^20^ Alfieri, N.L., Kusma, J.D., Heard-Garris, N. et al. Parental COVID-19 vaccine hesitancy for children: vulnerability in an urban hotspot. BMC Public Health 21, 1662 (2021). https://doi.org/10.1186/s12889-021-11725-5

^21^ He, K., Mack, W.J., Neely, M. et al. Parental Perspectives on Immunizations: Impact of the COVID-19 Pandemic on Childhood Vaccine Hesitancy. J Community Health 47, 39–52 (2022). https://doi.org/10.1007/s10900-021-01017-9

^22^ Fisher CB, Gray A, Sheck I. COVID-19 Pediatric Vaccine Hesitancy among Racially Diverse Parents in the United States. Vaccines. 2022; 10(1):31. https://doi.org/10.3390/vaccines10010031

^23^ David P, Fracci S, Wojtowicz J, et al. Ethnicity, Social Determinants of Health, and Pediatric Primary Care During the COVID-19 Pandemic. J Prim Care Community Health. 2022;13:21501319221112248. doi:10.1177/21501319221112248

^24^ Finney Rutten LJ, Zhu X, Leppin AL, et al. Evidence-Based Strategies for Clinical Organizations to Address COVID-19 Vaccine Hesitancy. Mayo Clin Proc. 2021;96(3):699-707. doi:10.1016/j.mayocp.2020.12.024

^25^ Khubchandani, J., Sharma, S., Price, J.H. et al. COVID-19 Vaccination Hesitancy in the United States: A Rapid National Assessment. J Community Health 46, 270–277 (2021). https://doi.org/10.1007/s10900-020-00958-x

^26^ Wiysonge CS, Ndwandwe D, Ryan J, et al. Vaccine hesitancy in the era of COVID-19: could lessons from the past help in divining the future?. Hum Vaccin Immunother. 2022;18(1):1-3. doi:10.1080/21645515.2021.1893062

^27^ Shah GH, Shankar P, Schwind JS, Sittaramane V. The Detrimental Impact of the COVID-19 Crisis on Health Equity and Social Determinants of Health. J Public Health Manag Pract. 2020;26(4):317-319. doi:10.1097/PHH.0000000000001200

^28^ Anderson MR, Hardy EJ, Battle CL. COVID-19 Vaccine Hesitancy during the Perinatal Period: Understanding Psychological and Cultural Factors to Improve Care and Address Racial/Ethnic Health Inequities. Women's Health Issues. 2022;32(4):317-321. doi:10.1016/j.whi.2022.04.001

^29^ Kiefer MK, Mehl R, Costantine MM, et al. Characteristics and perceptions associated with COVID-19 vaccination hesitancy among pregnant and postpartum individuals: A cross-sectional study. BJOG. 2022;129(8):1342-1351. doi:10.1111/1471-0528.17110

^30^ McGuire AL, Aulisio MP, Davis FD, et al. Ethical Challenges Arising in the COVID-19 Pandemic: An Overview from the Association of Bioethics Program Directors (ABPD) Task Force. Am J Bioeth. 2020;20(7):15-27. doi:10.1080/15265161.2020.1764138

^31^ Garg I, Hanif H, Javed N, Abbas R, Mirza S, Javaid MA, Pal S, Shekhar R, Sheikh AB. COVID-19 Vaccine Hesitancy in the LGBTQ+ Population: A Systematic Review. Infectious Disease Reports. 2021; 13(4):872-887. https://doi.org/10.3390/idr13040079

^32^ Newman PA, Reid L, Tepjan S, et al. COVID-19 vaccine hesitancy among marginalized populations in the U.S. and Canada: Protocol for a scoping review. PLoS One. 2022;17(3):e0266120. Published 2022 Mar 31. doi:10.1371/journal.pone.0266120

^33^ Oberg C, Hodges HR, Gander S, Nathawad R, Cutts D. The impact of COVID-19 on children's lives in the United States: Amplified inequities and a just path to recovery. Curr Probl Pediatr Adolesc Health Care. 2022;52(7):101181. doi:10.1016/j.cppeds.2022.101181

^34^ Salmon D, Opel DJ, Dudley MZ, Brewer J, Breiman R. Reflections On Governance, Communication, And Equity: Challenges And Opportunities In COVID-19 Vaccination. Health Aff (Millwood). 2021;40(3):419-425. doi:10.1377/hlthaff.2020.02254

^35^ Vicetti Miguel CP, Dasgupta-Tsinikas S, Lamb GS, Olarte L, Santos RP. Race, Ethnicity, and Health Disparities in US Children With COVID-19: A Review of the Evidence and Recommendations for the Future. J Pediatric Infect Dis Soc. 2022;11(Supplement_4):S132-S140. doi:10.1093/jpids/piac099

^36^ Cholera R, Falusi OO, Linton JM. Sheltering in Place in a Xenophobic Climate: COVID-19 and Children in Immigrant Families. Pediatrics. 2020;146(1):e20201094. doi:10.1542/peds.2020-1094

^37^ Flor LS, Friedman J, Spencer CN, et al. Quantifying the effects of the COVID-19 pandemic on gender equality on health, social, and economic indicators: a comprehensive review of data from March, 2020, to September, 2021. Lancet. 2022;399(10344):2381-2397. doi:10.1016/S0140-6736(22)00008-3

^38^ Abrams EM, Szefler SJ. COVID-19 and the impact of social determinants of health. Lancet Respir Med. 2020;8(7):659-661. doi:10.1016/S2213-2600(20)30234-4

^39^ Jones E, MacDougall H, Monnais L, Hanley J, Carstairs C. Beyond the COVID-19 crisis: Building on Lost Opportunities in the history of Public Health. FACETS. 2021Apr29;6(1):614–39.

^40^ Fronteira I, Sidat M, Magalhães JP, et al. The SARS-CoV-2 pandemic: A syndemic perspective. One Health. 2021;12:100228. doi:10.1016/j.onehlt.2021.100228

^41^ Rodrigues F, Block S, Sood S. What Determines Vaccine Hesitancy: Recommendations from Childhood Vaccine Hesitancy to Address COVID-19 Vaccine Hesitancy. Vaccines. 2022; 10(1):80. https://doi.org/10.3390/vaccines10010080

^42^ Kumar, Aditya (2022) Childhood Thriving in Urban and Rural Areas during COVID-19. Master Essay, University of Pittsburgh.

^43^ Parimi K, Gilkeson K, Creamer BA. COVID-19 vaccine hesitancy: Considerations for reluctance and improving vaccine uptake. Hum Vaccin Immunother. 2022;18(5):2062972. doi:10.1080/21645515.2022.2062972

^44^ Afolabi AA, Ilesanmi OS. Addressing COVID-19 vaccine hesitancy: Lessons from the role of community participation in previous vaccination programs. Health Promot Perspect. 2021;11(4):434-437. Published 2021 Dec 19. doi:10.34172/hpp.2021.54

^45^ Zhang Y, Fisk RJ. Barriers to vaccination for coronavirus disease 2019 (COVID-19) control: experience from the United States. Glob Health J. 2021;5(1):51-55. doi:10.1016/j.glohj.2021.02.005

^46^ Davenhall, W.F., Kinabrew, C. (2022). Geographic Information Systems in Health and Human Services. In: Kresse, W., Danko, D. (eds) Springer Handbook of Geographic Information. Springer Handbooks. Springer, Cham. https://doi.org/10.1007/978-3-030-53125-6_29

^47^ Afzal A, Shariff MA, Perez-Gutierrez V, Khalid A, Pili C, Pillai A, Venugopal U, Kasubhai M, Kanna B, Poole BD, Pickett BE, Redd DS, Menon V. Impact of Local and Demographic Factors on Early COVID-19 Vaccine Hesitancy among Health Care Workers in New York City Public Hospitals. Vaccines. 2022; 10(2):273. https://doi.org/10.3390/vaccines10020273

^48^ El-Mohandes A, White TM, Wyka K, et al. COVID-19 vaccine acceptance among adults in four major US metropolitan areas and nationwide. Sci Rep. 2021;11(1):21844. Published 2021 Nov 4. doi:10.1038/s41598-021-00794-6

^49^ Tharpe NL, McDaniel L. Using a Harm Reduction Model to Reduce Barriers to Vaccine Administration. J Midwifery Womens Health. 2021;66(3):308-321. doi:10.1111/jmwh.13259

^50^ Bloom DE, Cadarette D, Ferranna M. The Societal Value of Vaccination in the Age of COVID-19. Am J Public Health. 2021;111(6):1049-1054. doi:10.2105/AJPH.2020.306114

^51^ Isasi F, Naylor MD, Skorton D, Grabowski DC, Hernández S, Rice VM. Patients, Families, and Communities COVID-19 Impact Assessment: Lessons Learned and Compelling Needs. NAM Perspect. 2021;2021:10.31478/202111c. Published 2021 Nov 29. doi:10.31478/202111c

^52^ Hu M, Roberts JD, Azevedo GP, Milner D. The role of built and social environmental factors in COVID-19 transmission: A look at America’s capital city. Sustainable Cities and Society. 2021Feb;65:102580.

^53^ Wakeel F, Njoku A. Application of the Weathering Framework: Intersection of Racism, Stigma, and COVID-19 as a Stressful Life Event among African Americans. Healthcare. 2021; 9(2):145. https://doi.org/10.3390/healthcare9020145

^54^ Firebaugh, C.M., Beeson, T., Wojtyna, A., Bravo, L., Everson, T., Johnson, J. and Saldana, A. (2020) A Community Case Study on Geographic, Environmental, and Social Health Disparities in COVID-19 Disease: Yakima, Washington. Open Journal of Preventive Medicine, 10, 288-297. https://doi.org/10.4236/ojpm.2020.1011021

^55^ Rifat SAA, Liu W. One year into the pandemic: the impacts of social vulnerability on COVID-19 outcomes and urban-rural differences in the conterminous United States. Int J Environ Health Res. 2022;32(12):2601-2619. doi:10.1080/09603123.2021.1979196

^56^ Mah JC, Kulkarni A, Forman R, Mossialos E. Social and Physical Environment Disparities Contribute to Mortality Outcomes in COVID-19 Pandemic in the United States. Journal of Health Policy and Economics. 2021;1(1):.

^57^ Magnan S. Social Determinants of Health 201 for Health Care: Plan, Do, Study, Act. NAM Perspect. 2021;2021:10.31478/202106c. Published 2021 Jun 21. doi:10.31478/202106c

^58^ Bogan E, Adams‐Bass VN, Francis LA, Gaylord‐Harden NK, Seaton EK, Scott JC, et al. “wearing a mask won't protect us from our history”: The impact of covid‐19 on Black Children and families. Social Policy Report. 2022Jul18;35(2):1–33.

^59^ Cole M, Jolliffe M, So-Armah C, Gottlieb B. Power and participation: How community health centers address the determinants of the social determinants of health. NEJM Catalyst. 2022Jan1;3(1).

^60^ Gil RM, Freeman TL, Mathew T, Kullar R, Fekete T, Ovalle A, et al. Lesbian, gay, bisexual, transgender, and queer (LGBTQ+) communities and the coronavirus disease 2019 pandemic: A call to break the cycle of structural barriers. The Journal of Infectious Diseases. 2021Jul29;224(11):1810–20.

^61^ Brakefield, Whitney, "Design and Development of the Urban Population Health Observatory to Improve Disease Surveillance and Response." Ph.D. diss., University of Tennessee, 2022. https://trace.tennessee.edu/utk_graddiss/7153

^62^ Loukaitou-Sideris, A., Wasserman, J. L, Caro, R., & Ding, H. (2021). Homelessness in Transit Environments Volume II: Transit Agency Strategies and Responses. UCLA: Institute of Transportation Studies. Retrieved from https://escholarship.org/uc/item/87b0v8cr

^63^ Perrigo JL, Samek A, Hurlburt M. Minority and low-SES families' experiences during the early phases of the COVID-19 pandemic crisis: A qualitative study. Child Youth Serv Rev. 2022;140:106594. doi:10.1016/j.childyouth.2022.106594

^64^ Harrison, Teresa; Pardo, Theresa; Carleo-Evangelist, Jordan; and Warner, Lynn. 6-24-2021. "Minority Health Disparities in a 21st-century Pandemic: A Comprehensive Report of Project Research Focused on New York" Understanding and eliminating minority health disparities in a 21st-century pandemic: A White Paper Collection. University at Albany, SUNY: Scholars Archive. https://scholarsarchive.library.albany.edu/covid_mhd_nys_white_papers/11

^65^ Chumpa, Hamida, "Exploring Social Determinants of COVID-19 related Sickness and Suffering in the Bronx" (2021). CUNY Academic Works. https://academicworks.cuny.edu/bb_etds/112

^66^ Bonotti, M., Zech, S.T. (2021). The Human, Economic, Social, and Political Costs of COVID-19. In: Recovering Civility during COVID-19. Palgrave Macmillan, Singapore. https://doi.org/10.1007/978-981-33-6706-7_1

^67^ Chen, Y., Jiao, J., Bai, S., & Lindquist, J. (2020). Modeling the Spatial Factors of COVID-19 in New York City. SSRN Electronic Journal. http://dx.doi.org/10.2139/ssrn.3606719

^68^ Byttebier, K. (2022). Covid-19 and Inequality. In: Covid-19 and Capitalism. Economic and Financial Law & Policy – Shifting Insights & Values, vol 7. Springer, Cham. https://doi.org/10.1007/978-3-030-92901-5_10

^69^ Barzilay R, Moore TM, Greenberg DM, et al. Resilience, COVID-19-related stress, anxiety and depression during the pandemic in a large population enriched for healthcare providers. Transl Psychiatry. 2020;10(1):291. Published 2020 Aug 20. doi:10.1038/s41398-020-00982-4

^70^ Bavel, J.J.V., Baicker, K., Boggio, P.S. et al. Using social and behavioral science to support COVID-19 pandemic response. Nat Hum Behav 4, 460–471 (2020). https://doi.org/10.1038/s41562-020-0884-z

^71^ Carrión D, Colicino E, Pedretti NF, et al. Neighborhood-level disparities and subway utilization during the COVID-19 pandemic in New York City. Nat Commun. 2021;12(1):3692. Published 2021 Jun 17. doi:10.1038/s41467-021-24088-7

^72^ K C M, Oral E, Straif-Bourgeois S, Rung AL, Peters ES. The effect of area deprivation on COVID-19 risk in Louisiana. PLoS One. 2020;15(12):e0243028. Published 2020 Dec 3. doi:10.1371/journal.pone.0243028

^73^ Anderson KF, Ray-Warren D. Racial-Ethnic Residential Clustering and Early COVID-19 Vaccine Allocations in Five Urban Texas Counties. J Health Soc Behav. 2022;63(4):472-490. doi:10.1177/00221465221074915

^74^ Benninger E, Schmidt-Sane M, Spilsbury JC. Conceptualizing Social Determinants of Neighborhood Health through a Youth Lens. Child Indic Res. 2021;14(6):2393-2416. doi:10.1007/s12187-021-09849-6

^75^ Bilal U, Mullachery PH, Schnake-Mahl A, et al. Heterogeneity in Spatial Inequities in COVID-19 Vaccination Across 16 Large US Cities. Am J Epidemiol. 2022;191(9):1546-1556. doi:10.1093/aje/kwac076

^76^ Choi K, Romero R, Guha P, et al. Community Health Worker Perspectives on Engaging Unhoused Peer Ambassadors for COVID-19 Vaccine Outreach in Homeless Encampments and Shelters. J Gen Intern Med. 2022;37(8):2026-2032. doi:10.1007/s11606-022-07563-9

^77^ Crist, K., Benmarhnia, T., Frank, L.D. et al. The TROLLEY Study: assessing travel, health, and equity impacts of a new light rail transit investment during the COVID-19 pandemic. BMC Public Health 22, 1475 (2022). https://doi.org/10.1186/s12889-022-13834-1

^78^ DiRago NV, Li M, Tom T, et al. COVID-19 Vaccine Rollouts and the Reproduction of Urban Spatial Inequality: Disparities Within Large US Cities in March and April 2021 by Racial/Ethnic and Socioeconomic Composition. J Urban Health. 2022;99(2):191-207. doi:10.1007/s11524-021-00589-0

^79^ Ezell JM, Griswold D, Chase EC, Carver E. The blueprint of disaster: COVID-19, the Flint water crisis, and unequal ecological impacts. The Lancet Planetary Health. 2021May;5(5).

^80^ Hillis SD, Blenkinsop A, Villaveces A, et al. COVID-19-Associated Orphanhood and Caregiver Death in the United States [published online ahead of print, 2021 Oct 7]. Pediatrics. 2021;e2021053760. doi:10.1542/peds.2021-053760

^81^ Mollalo A, Tatar M. Spatial Modeling of COVID-19 Vaccine Hesitancy in the United States. Int J Environ Res Public Health. 2021;18(18):9488. Published 2021 Sep 8. doi:10.3390/ijerph18189488

^82^ Morgan KM, Maglalang DD, Monnig MA, Ahluwalia JS, Avila JC, Sokolovsky AW. Medical Mistrust, Perceived Discrimination, and Race: a Longitudinal Analysis of Predictors of COVID-19 Vaccine Hesitancy in US Adults [published online ahead of print, 2022 Aug 1]. J Racial Ethn Health Disparities. 2022;1-10. doi:10.1007/s40615-022-01368-6

^83^ Ransome Y, Ojikutu BO, Buchanan M, Johnston D, Kawachi I. Neighborhood Social Cohesion and Inequalities in COVID-19 Diagnosis Rates by Area-Level Black/African American Racial Composition. J Urban Health. 2021;98(2):222-232. doi:10.1007/s11524-021-00532-3

^84^ Sacarny A, Daw JR. Inequities in COVID-19 Vaccination Rates in the 9 Largest US Cities. JAMA Health Forum. 2021;2(9):e212415. doi:10.1001/jamahealthforum.2021.2415

^85^ Xiao W, Wei YD, Wu Y. Neighborhood, built environment and resilience in transportation during the COVID-19 pandemic. Transp Res D Transp Environ. 2022;110:103428. doi:10.1016/j.trd.2022.103428

^86^ Zeng C, Zhang J, Li Z, et al. Population Mobility and Aging Accelerate the Transmission of Coronavirus Disease 2019 in the Deep South: A County-Level Longitudinal Analysis. Clin Infect Dis. 2022;74(Suppl_3):e1-e3. doi:10.1093/cid/ciac050

^87^ Gin JL, Balut MD, Alenkin NR, Dobalian A. Responding to COVID-19 While Serving Veterans Experiencing Homelessness: The Pandemic Experiences of Healthcare and Housing Providers. J Prim Care Community Health. 2022;13:21501319221112585. doi:10.1177/21501319221112585
